# Three-dimensional printing of lifelike PET phantoms

**DOI:** 10.64898/2026.05.11.26352857

**Authors:** Yinglin Ge, Elizabeth J Li, Stephen McDonald, Michael Geagan, Michael J Parma, Min Gao, Kai Mei, Pouyan Pasyar, Jessica Im, Florence M Muller, Austin R Pantel, Joel S Karp, Peter B Noël

**Affiliations:** Department of Radiology, Perelman School of Medicine, University of Pennsylvania, Philadelphia, USA; Department of Bioengineering, University of Pennsylvania, Philadelphia, USA; Faculty of Engineering and Architecture, Ghent University, Ghent, Belgium

## Abstract

**Background:** Realistic PET/CT phantoms are essential for system evaluation, protocol optimization, and validation of advanced reconstruction methods. However, existing phantoms are often limited by simplified geometries, spatially uniform activity patterns, and complex preparation procedures.

**Purpose:** To develop and evaluate PixelPrint^PET^, a 3D printing-based method for fabricating anatomically realistic PET/CT phantoms with spatially heterogeneous radiotracer distributions and a single-solution filling workflow that avoids physical compartmentalization.

**Methods:** PixelPrint^PET^ generates voxel-based printing instructions that encode spatially varying infill, which is realized during printing through modulation of filament extrusion, enabling heterogeneous activity distributions without compartmentalization of radioactivity at different activity concentrations. Calibration phantoms and anatomically structured phantoms were designed and printed using high-flow polylactic acid (PLA), with anatomical inputs derived from either digital atlas-based models or patient imaging data. The printed phantoms were subsequently filled by immersion in a radioactive solution, allowing activity distribution to be controlled by the internal porous structure. A bottom-up filling procedure with reduced surface tension was developed to ensure uniform infiltration and minimize air entrapment. Phantoms were imaged on the PennPET Explorer PET/CT system, and quantitative performance was evaluated using contrast recovery coefficient (CRC), target-to-background ratio (TBR), and comparisons with simulated or patient-derived reference data.

**Results:** A strong linear relationship between infill ratio and normalized signal (R^2^ = 0.998) was demonstrated by the calibration phantom, enabling reliable mapping between structure and activity. Additionally, air entrapment was minimized to less than 1% of the total phantom volume. In the contrast recovery phantom, CRC values were consistent with measurements using traditional phantoms. The brain phantom reproduced atlas-derived contrast patterns, with gray-to-white matter differences within 5% after accounting for resolution and other system effects. The patient-based thorax phantom showed high reproducibility across repeated scans, with differences within 3%, and closely matched the input patient image with regional differences within 10% in all regions except the lung.

**Conclusions:** PixelPrint^PET^ enables the fabrication of realistic, reproducible, and versatile PET/CT phantoms with a voxel-level control of the activity distribution. This approach provides a practical solution for generating patient-specific and application-specific phantoms, with the potential to accelerate system validation, protocol development, and clinical translation of advanced PET/CT technologies.

## 1. INTRODUCTION

The combination of positron emission tomography and computed tomography (PET/CT) allows for the integration of molecular imaging from PET with detailed anatomical information from CT, providing greater diagnostic sensitivity and specificity than either modality alone. In addition to serving as an anatomical reference, CT data is routinely used for attenuation correction in PET, which is needed for quantitative accuracy.^1^ The PET/CT scan with fluorodeoxyglucose (FDG) as radiotracer has become a standard tool in diagnosing cancer, cardiovascular diseases, and neurological disorders.^2,3^

Traditional phantoms, such as the NEMA Image Quality phantom, are widely used for scanner performance characterization^4^. Other designs, including the ACR PET phantom and the Hoffman 3D Brain phantom, support multicenter studies, accreditation efforts, and protocol standardization.^5^ While providing a foundation for performing standardized measurements, these phantoms commonly used in PET/CT are limited in their correspondence to anatomic features. Notably, lesions in the NEMA and ACR PET phantoms are glass or plastic spheres with walls, whose thickness (typically ∼ 1 mm) has a measurable impact on quantifying the uptake for small lesions (i.e., 1 cm diameter or less). The Hoffman 3D Brain phantom is based on a human MRI and mimics the fine structure anatomy of the brain in the transverse plane, but only coarsely in the axial planes. In addition, conventional phantom preparation often requires carefully measured activity solutions to generate a specified contrast in the structures, requiring technical expertise and experience handling radioactive materials to produce standardized measurements.

The development of anatomically accurate imaging phantoms that replicate specified radiotracer distribution and X-ray attenuation is essential for advancing standardized measurements for PET/CT studies. Recent studies have explored the use of 3D printing to fabricate more customizable phantoms. Approaches such as printing FDG-containing ink on paper sheets followed by stacking can produce three-dimensional activity distributions, ^6,7^ while other methods have incorporated radioactive materials directly into printed structures.^8^ Although these techniques enable anatomically shaped phantoms, they are constrained by short isotopic half-lives, limited activity flexibility, and challenges in storage and reuse. Alternative approaches based on printing hollow shells derived from patient segmentations can reproduce organ shapes and activity patterns,^9–11^ but often require complex preparation and may introduce boundary artifacts and partial volume effects that limit the representation of fine structures. In addition, these methods are typically restricted to a limited number of discrete activity levels, which does not reflect the continuous variation in tracer uptake observed in the human body. More advanced methods, such as high-resolution 3D printing phantom combined with list–mode– based post-processing for adjustable contrast, have achieved greater structural detail and varied contrast without introducing confounding boundary effects from enclosing walls, but are typically restricted to simplified contrasts such as gray and white matter and may involve substantial computational complexity.^12^ Overall, it remains challenging to simultaneously achieve anatomical realism, spatially heterogeneous activity distributions, and practical fabrication workflows.

We recently introduced PixelPrint to manufacture CT phantoms with accurate attenuation profiles and textures that mimic human features.^13,14^ This 3D printing technique creates voxel-based printer instructions by modeling density as a ratio of filament to voxel volume, thereby generating sub-resolution partial volume effects to replicate realistic tissue densities and textures. In this work, we introduce technology to manufacture PET phantoms, referred to as PixelPrint^PET^, a method for fabricating phantoms with specified effective radiotracer activity distributions.^15^ This approach enables the generation of phantoms with anatomically realistic structures and heterogeneous activity distributions, advancing the development of more representative imaging models for clinical translation.

The proposed method is designed to overcome key limitations of existing approaches by eliminating physical boundaries between regions, enabling representation of a full range of tissue structures, including small features, and supporting spatially heterogeneous activity distributions with realistic image texture. In addition, PixelPrint^PET^ allows flexible input from either patient images without segmentation or digital models, while maintaining a simple workflow for phantom filling with reproducible results for radiotracer uptake. In this work, we present the first results of a brain phantom (using a high-resolution atlas model as input) and a torso phantom (using a patient image as input) to demonstrate the potential of this new methodology.

## 2. METHODS

### 2.1 3D-printing

In PET imaging, contrast arises from differences in radiotracer uptake across spatial locations. To replicate this, our PixelPrint^PET^ phantom employed varying infill ratios, defined as the proportion of filament occupying a given unit of volume. The remaining space is thus air, or when submerged, radiotracer solution. The remaining space is thus air, or when submerged, radiotracer solution. To achieve variable line infill ratios, the infill ratio was controlled by adjusting the printing line width as the printing line distance was fixed at 2.0 mm, and the printed layer height at 0.2 mm:

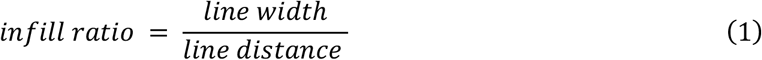

The higher the infill ratio, the lower the PET signal. By adjusting the printed line width, infill ratios from 0 to 100% can be theoretically achieved. In practice, the usable infill range is limited by the minimum line width required to maintain structural integrity and the maximum infill level beyond which air bubbles are likely to become trapped. From our previous study, the minimum infill ratio required to maintain structural stability was 5%, while the maximum infill ratio that avoided noticeable air trapping was 95%.^15^

We used the PixelPrint software^13,16^ to design the phantoms and to generate G-code, the machine language that specifies 3D printing instructions. Based on the generated G-code, the phantoms were printed using a Fused Deposition Modeling (FDM) 3D printer (Prusa XL 3D printer, Prusa Research a.s., Prague, Czech Republic) equipped with a 0.4 mm brass nozzle and high-flow polylactic acid (PLA) filament with a diameter of 1.75 mm (Protopasta Filament, Protoplant Inc., Vancouver, WA, USA). We used a commercial high-flow PLA as the printing material because PLA provides low positron attenuation similar to water. In addition, the material contains a wetting agent that reduces surface tension within the printed structure, which lowers the likelihood of trapped air bubbles during radiotracer solution filling. To improve the filament-to-bed adhesion, the build plate was heated to 50 °C.

#### 2.1.2 Phantom design

##### A. Technical phantoms

Two technical phantoms were designed to establish a baseline for subsequent development. To generate different activity concentration levels in the PET images, the infill ratio was varied to create fillable voxels with different effective volumes. The printed line widths were kept smaller than the spatial resolution of the PennPET Explorer system (4 mm) so that individual filament lines would not be visible in the reconstructed images. Both phantoms were printed with a height of 40 mm to minimize edge-related partial-volume effects (PVE) at the center of the phantom.

First, a single cuboid **infill calibration phantom** with dimensions of 150 mm in width was designed to get the relationship between infill ratio and activity level. The phantom was divided into 25 square sections, each measuring 30 × 30 × 40 mm. Each section was printed with a distinct infill ratio ranging from 5% to 100% in increments of 5%, with intentional overlap of selected ratios to enable repeatability checks (Figure 1).

**Figure 1.**
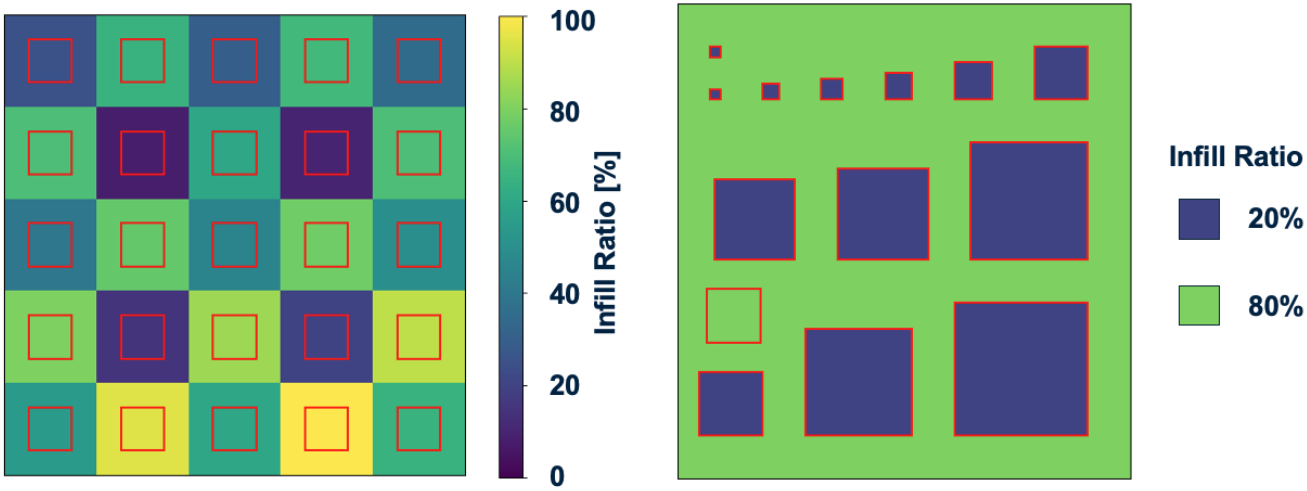
Schematic of the quality control phantoms and regions of interest (ROIs, red boxes) used for signal measurement and contrast calculation. Red lines contour the ROI regions.

Secondly, a **contrast recovery phantom** with multiple region sizes was designed to evaluate size-dependent contrast recovery and PVE of PixelPrint^PET^. The infill ratios were set to 20% for the hot regions and 80% for the background, resulting in an activity concentration ratio between hot and background regions based on the infill ratio. Thirteen square hot regions with side lengths ranging from 4 to 50 mm were arranged with a side-to-side spacing of 15 mm, Figure 2B. This design spans a range of feature sizes for assessing contrast recovery across spatial scales.

**Figure 2.**
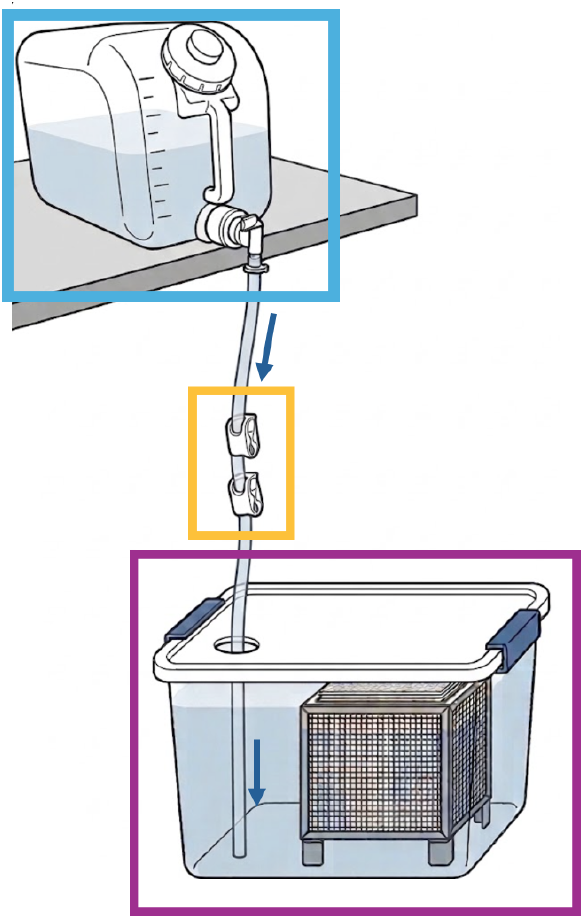
Experiment setup. The FDG solution with wetting agent was mixed in the carboy (blue box) and flowed (direction shown by blue arrow) through the tube into the bottom of the phantom container (purple box). The phantom was placed on a customized 3D printed holder. The flow rate was controlled by the valve of the bottom outlet of the carboy and the pinch clip fluid flow control valve (yellow box).

##### B. Brain phantom

The PET brain phantom was created using the BigBrain MR dataset,^17^ which provides a digital brain model with realistic anatomical detail and segmentations up to 100 μm resolution. Based on these labels and available FDG data,^18,19^ we assigned regional activity concentration ratios to generate a PET contrast map.^20^ The ratios were normalized and scaled to span from 1.0 in cerebrospinal fluid (CSF) to 8.0 in the subthalamic and red nuclei, and with a gray matter (GM) to white matter (WM) contrast of 3:1, reflecting a realistic range of contrast ratios. Using the calibration phantom results, each labeled region was then mapped to a corresponding infill ratio. Two identical brain phantoms, each with dimensions of approximately 18 × 19 × 14 cm^3^, were fabricated to evaluate degassing efficiency and contrast repeatability.

##### C. Thorax phantom

To evaluate the feasibility of generating patient-specific phantoms, a clinical PET dataset was used as the direct input for phantom fabrication. This study was approved by the University of Pennsylvania Institutional Review Board and performed under IRB 843546. Written informed consent was obtained. A total-body PET scan from a healthy female subject (BMI: 21.2 kg/m^2^, tracer: ^18^F-FDG, activity: 370 MBq) was acquired on the PennPET Explorer PET/CT following normal protocol. The PET image data at 50-60 min p.i. used in this study was collected retrospectively and anonymized.

An 80 mm axial range was extracted to cover the region from the aorta arch to the superior portion of the liver. Background was removed by cropping, and voxels with very low activity, defined as the lowest 2% of intensity values, were excluded to suppress noise-dominated signals. The remaining voxel intensities were normalized to the minimum retained value and then converted to infill ratio using the calibration results. Notches were printed along X and Y axes of the torso phantom as fiducial markers for spatial alignment with the patient images.

### 2.2 Experimental setup

To fill the phantom with radioactive solution while minimizing air entrapment, a simple and robust filling procedure was developed. Approximately 180 MBq of ^18^F-FDG was mixed with water in a carboy to achieve the desired activity concentration. A small amount of Tween 20 (polysorbate 20, Thermo Fisher Scientific, MA, USA) was added at a final concentration of 0.1% (v/v) to reduce surface tension and facilitate filling. The phantom was placed in a container using customized 3D-printed PLA holders that kept it level and elevated slightly above the container base, allowing space beneath the phantom for fluid entry. Tubing connected to the carboy was positioned so that its outlet remained at the bottom of the filling container, ensuring that the incoming solution stayed submerged and did not introduce air bubbles. Plastic pinch clip fluid flow control valves were used to maintain a low, steady flow, while a height difference between the carboy and the container enabled gravity-driven delivery of the solution into the container. This setup allowed the solution to gradually infiltrate the phantom through capillary action, while hydrostatic equilibrium ensured uniform filling and minimized air entrapment. Because the required fill volume varied across phantoms, the flow rate and total filling time were adjusted as needed. Filling was continued until the liquid level reached approximately 1 to 2 cm above the phantom surface.

After completion of the filling procedure, the sealed container was transferred to the scanner bed. Imaging was performed on a PET/CT system at the University of Pennsylvania, consisting of a clinical dual-layer CT scanner (IQon Spectral CT, Philips Healthcare, The Netherlands)^21^ and the PennPET Explorer, ^22^ a whole-body PET system with a 142 cm axial field of view. All the scans were conducted under normal PET/CT protocol. CT images were acquired at 120 kVp and 11 mAs and reconstructed with a voxel size of 1.17 × 1.17 × 1.25 mm^3^. Following verification of air bubble removal and proper phantom positioning, PET data were acquired for 30 minutes. A second 30-minute scan was performed after a 90-minute delay to determine that the activity distribution is stable and to assess measurement consistency for brain and thorax phantoms. The PET images were reconstructed into 2×2×2mm^3^ voxel size using list-mode time-of-flight OSEM (5 iterations, 25 subsets) with CT-based attenuation correction and scatter correction.^23^

### 2.3 Analysis

#### 2.3.1 Filling validation

CT images were used to evaluate the effectiveness of the filling process. A reference mask was generated from the printing instruction file to represent the target geometry, and CT images were registered to this reference. The total phantom volume was calculated from the mask. The phantom region was then extracted based on the mask, and the corresponding Hounsfield Unit (HU) values were extracted from the CT images.

Voxels that were fully filled with solution and PLA exhibited positive HU values, whereas voxels containing trapped air showed values between −1000 and 0 due to partial volume effects. The residual air volume was estimated by calculating the fraction of voxels within this HU range relative to the total phantom volume.

#### 2.3.2 Contrast measurement

For the infill calibration phantom, regions of interest (ROIs) with a fixed size of 14 × 14 mm^2^ (around half of the region size) were manually placed on the PET images on five consecutive central slices. Mean activity concentrations and standard deviations were measured for each infill ratio. Linear regression was performed to evaluate the relationship between the normalized signal and infill ratio, and Pearson’s correlation coefficient was calculated. This relationship was used to establish a calibration curve for phantom design.

To assess the contrast recovery vs. region size, ROIs matching the size and shape of the designed hot regions were placed on the central five slices of the contrast recovery phantom, along with a 10 × 10 mm^2^ ROI in the background region at least 1 cm from the hot regions. Mean and standard deviation of activity concentration were measured, and the contrast between hot and background regions was calculated for each feature size. The results were quantified using the contrast recovery coefficient (CRC):

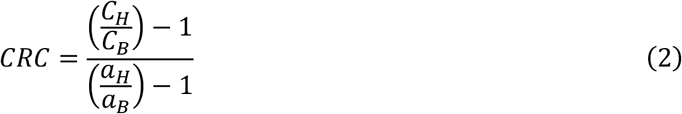

where *C*_*H*_ is the average counts in the hot ROI, and *C*_*B*_ is the average counts of the background ROI, while *a*_*H*_ and *a*_*B*_ are the activity concentrations in the hot sphere and the background, respectively.^24^

For the brain phantom, the same input dataset was simulated using GATE ^25^ with the same scanner geometry, detector design and reconstruction method as used in the phantom image acquisition. The simulation was designed to provide a reference image representing the expected phantom signal on the PennPET Explorer under ideal correction conditions, while preserving the effects of system geometry and spatial resolution. Attenuation and scatter effects were not explicitly included in the simulation; rather, we assume that accurate attenuation and scatter corrections are available, as in standard PET image reconstruction. Normalization correction was applied to account for detector sensitivity variations and to produce images representative of the system response. ^20^

Quantitative analysis was performed on both the simulated and phantom images. Region boundaries were defined directly from the labeled map used to generate the input data, which delineated regions with different contrasts. After image registration using SimpleITK in Python, mean activity concentrations were calculated for each region using the central 20 slices. The target-to-background ratio (TBR), with white matter as the reference background, was used as the evaluation metric. Differences between the simulated and measured phantom images were then quantified to assess agreement.

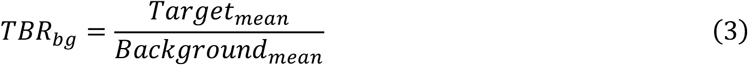

For the thorax phantom, which was derived from a patient image with normalized SUV values, the phantom and patient images were first registered. ROIs with volumes of at least 1 cm^3^ were placed in selected anatomical regions, and TBR_aorta_ with the aorta blood pool as reference background was then generated.

In addition, line profiles were extracted from corresponding locations in patient and phantom images and normalized to the mean aorta signal to enable direct comparison.

## 3. RESULTS

All phantoms were filled with FDG solution, with trapped air accounting for less than 1% of the total phantom volumes validated from CT.

For the infill calibration phantom, the mean values measured from regions of interest corresponding to infill ratios between 5% and 95% were normalized to the background region consisting of pure FDG solution and plotted in Figure 3A. A strong linear relationship was observed between infill ratio and normalized signal, with a Pearson correlation coefficient of R^2^ = 0.998. This relationship enables the use of the infill ratio as a reliable surrogate for normalized activity concentration and provides a basis for subsequent calibration. The contrast between different infill levels can also be derived from this calibration curve.

**Figure 3.**
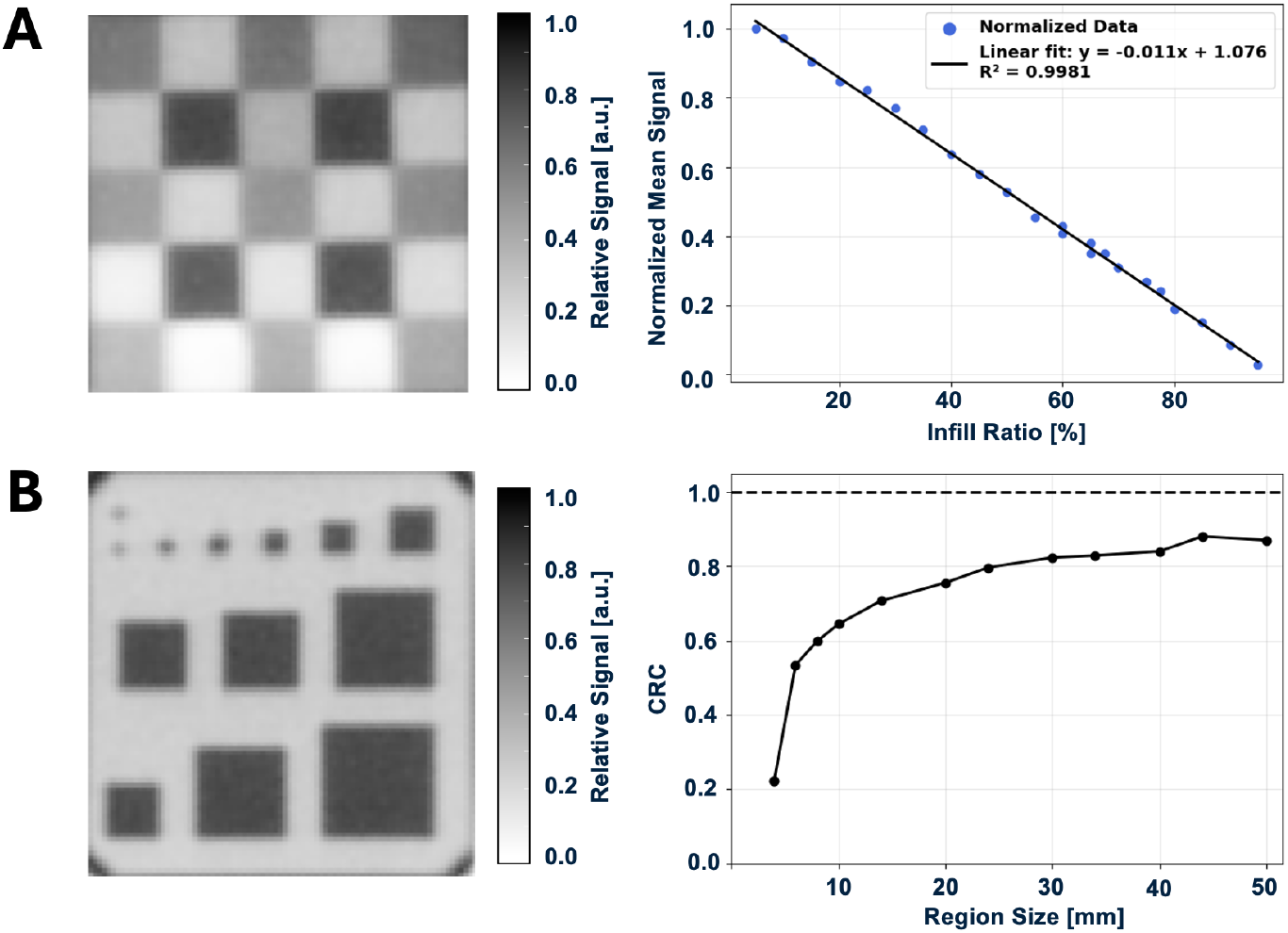
(A) Infill calibration phantom. Left: PET image of the central slice. Right: Linear regression between infill ratio and normalized signal. Blue markers represent mean values measured from cuboid ROIs placed in the central slice. The black line indicates the linear fit, and R^2^ denotes the coefficient of determination. (B) Contrast recovery phantom. Left: PET image of the central slice. Right: CRC as a function of hot region size.

Based on the calibration curve, the selected infill ratios of the contrast recovery phantom (20% for the high-signal region and 80% for the background) correspond to an expected contrast of 4.25. The measured contrast recovery coefficients are shown in Figure 3B. The recovered contrast approached a plateau for feature sizes larger than 44 mm, consistent with measurements obtained using traditional phantoms, such as the NEMA IQ with fillable glass or plastic spheres in a warm background^22^. While the CRC drops off rapidly below a region size of 10-mm, quantitative measurement of a 4-mm region is limited given the PET scanner’s spatial resolution (4-mm) and the image voxel size (2-mm), but we note that measurements of the two structures were consistent with each other, with a difference of less than 0.02, demonstrating the consistency of defining even small structures with this methodology.

The PixelPrint^PET^ brain phantoms were fabricated successfully without structural failure, and the reconstructed PET images closely resembled the simulated images from GATE (Figure 4). For quantitative evaluation, white matter was selected as the reference background due to its relatively large and homogeneous regions, which reduce the impact of partial volume effects and enable more stable measurements. Distinct differences in TBR_WM_ across regions demonstrate that the phantom successfully reproduces regional contrast, enabling clear separation of anatomical structures. Quantitatively, the measured TBR_WM_ values agree well with the simulation for most regions, with differences within 10%, while a larger deviation was observed for CSF (Figure 5). TBR_WM_ measurements showed good reproducibility across repeated scans and independently fabricated phantoms, with low variability.

**Figure 4.**
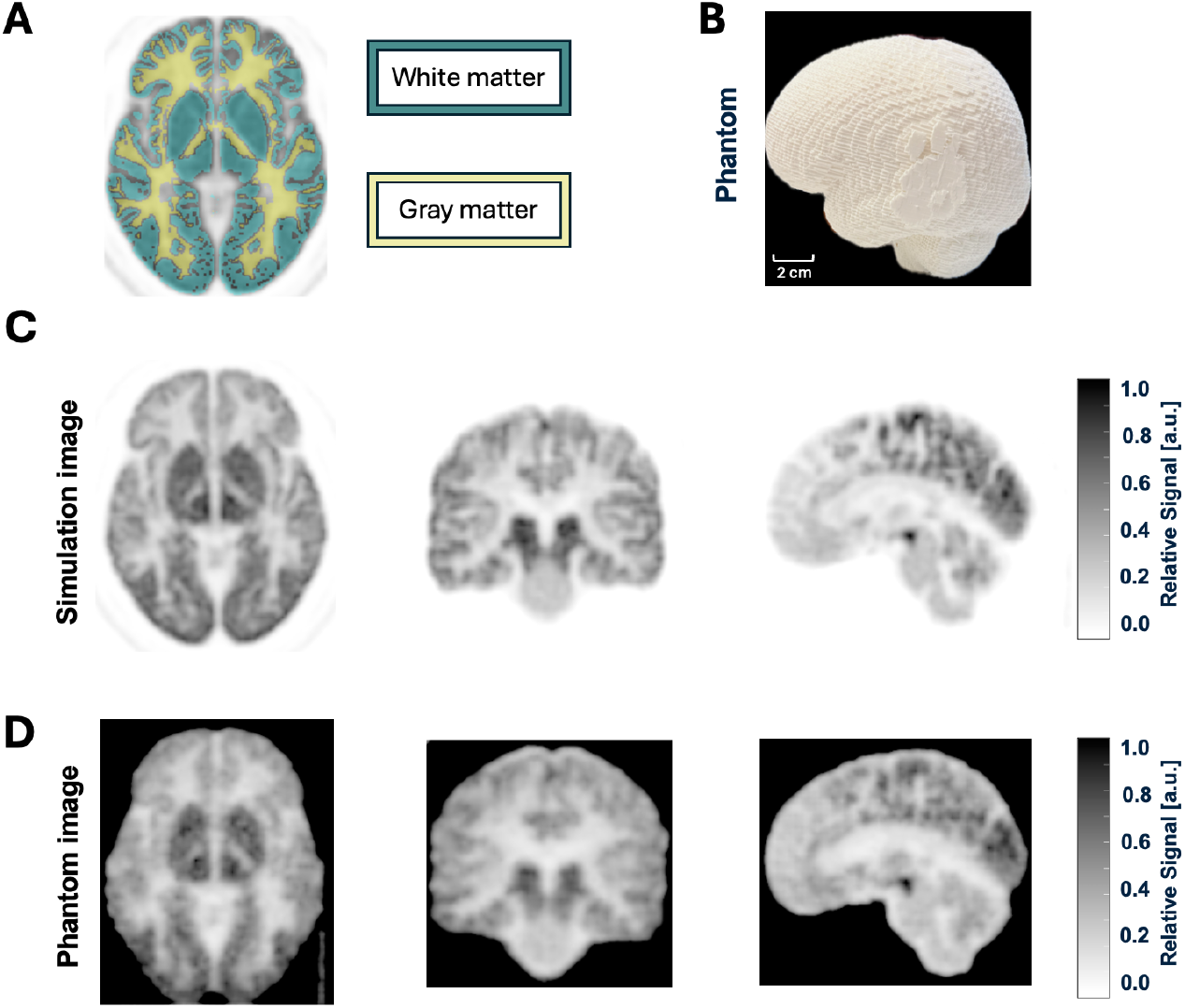
Whole-brain phantom. (A) Labeled maps from the BigBrain atlas were used both as input for phantom design and as reference maps for region-based analysis, providing spatial definitions of anatomical regions for quantitative evaluation. Here, white matter (cyan) and gray matter (yellow) labels, as examples, were overlaid on the results of the GATE simulation. (B) photograph of the 3D-printed brain phantom with scale bar. (C) Images of the phantom from the GATE simulation (C) were visually similar to those of the 3D-printed results (D).

**Figure 5.**
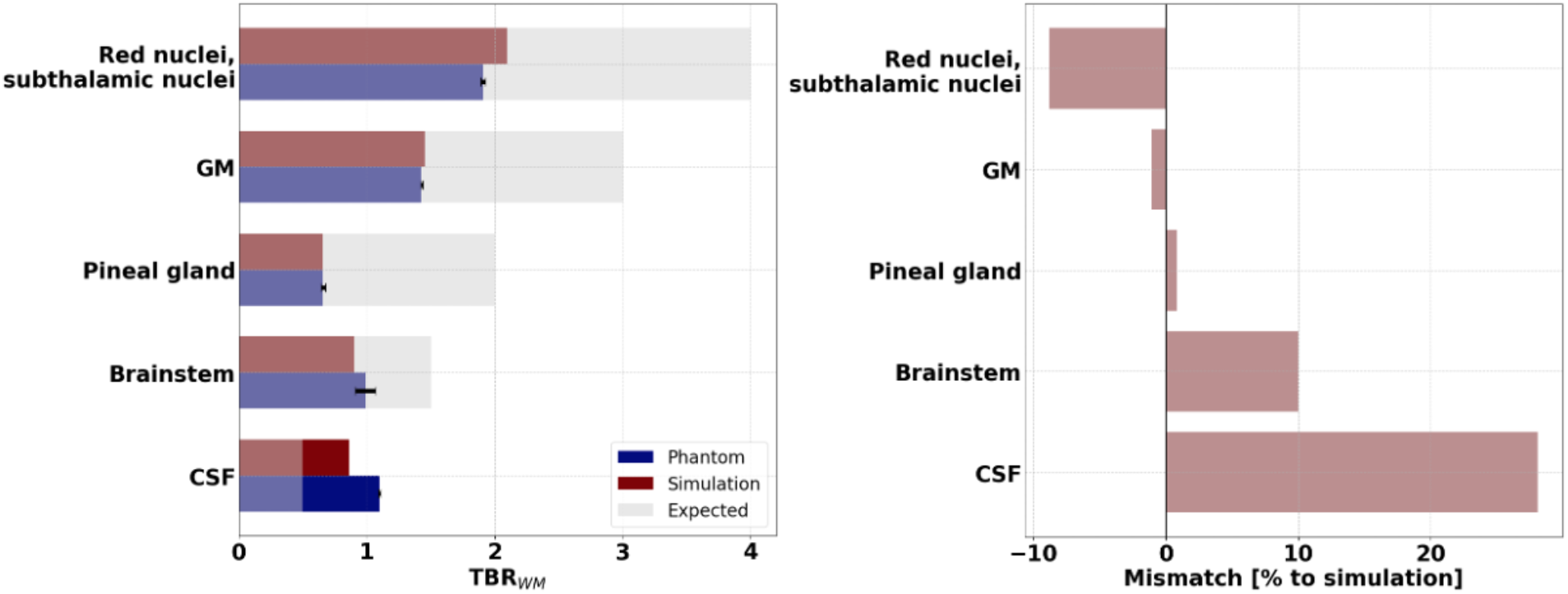
(Left) The TBR_WM_ of the measured phantom (blue), simulated (red) image and expected values (gray). The expected values were defined based on the assigned activity concentration ratios from the input atlas. Error bars were shown for the phantom images, indicating the standard deviation between two identical phantoms and two scans. (Right) Mismatch between measured phantom and simulation values, with simulation values as reference.

The thorax phantom was generated based on voxel-wise normalized SUV values, and its PET images were directly compared with the input patient data. As shown in Figure 6, the phantom images closely resemble the patient images, with clear contrast between organs and anatomical regions. The TBR_aorta_ is summarized in Table 1. All evaluated regions, except for the lungs, agreed with the patient data within 10%. Measurements across repeated scans were highly consistent, with differences below 3%, indicating stable performance across different count levels.

**Figure 6.**
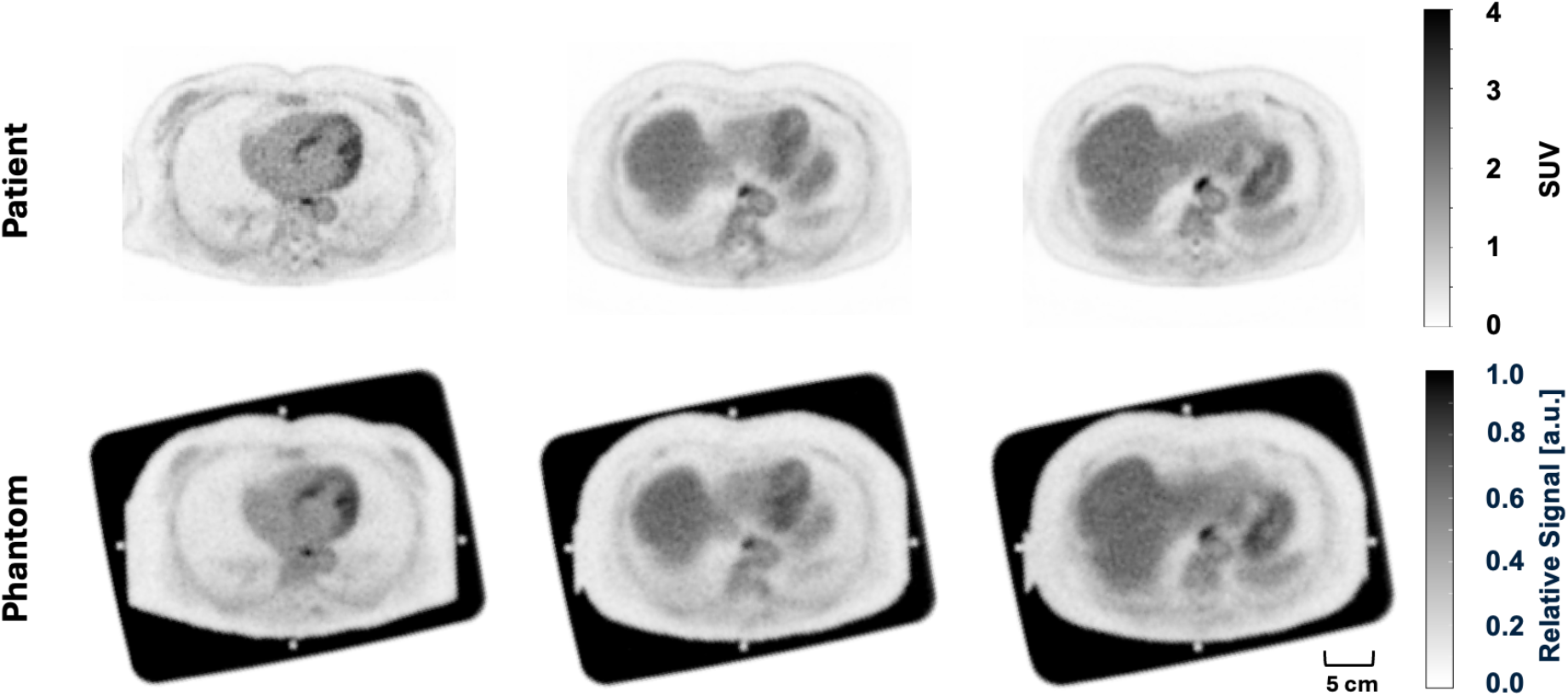
Representative slices from the patient and corresponding thorax phantom images. The scale bar is shown below the phantom image.

**Table 1.** Results of TBR_aorta_ for patient and phantom image.

| Structure | ROI size (cm<sup>3</sup>) | Patient $TBR_{aorta}$ | Phantom $TBR_{aorta}$ | Error (% to patient) |
| --- | --- | --- | --- | --- |
| Left ventricle | 6.1 | 1.01 | 0.99 | +3.00 |
| Liver | 4.6 | 1.47 | 1.44 | +1.62 |
| Myocardium | 2.0 | 1.76 | 1.63 | -6.80 |
| Vertebra | 1.6 | 1.22 | 1.17 | -4.57 |
| Lung | 4.0 | 0.22 | 0.30 | +35.4 |

Line profiles normalized to the mean activity in the aorta are shown in Figure 7. The line profile of the PixelPrint^PET^ patient-based phantom closely matches that of the original patient image, demonstrating preserved spatial signal transitions. Overall, the phantom reproduces both the contrast and spatial variation observed in the patient data.

**Figure 7.**
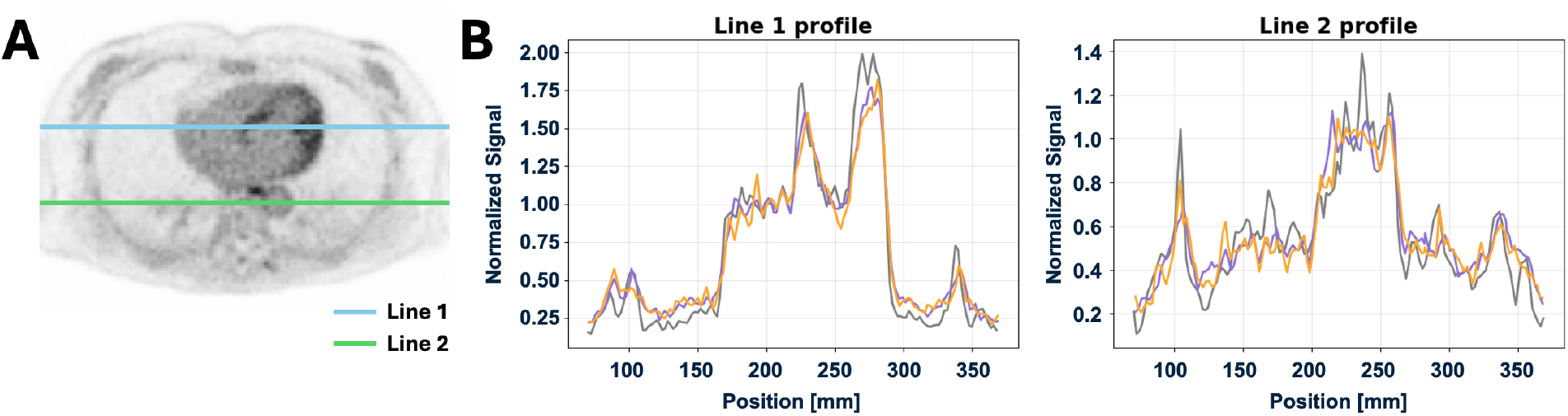
Line profile analysis of the thorax phantom. (A) Locations of the two-line profiles overlaid on the patient and phantom images. (B) Line profiles of measured SUVs, normalized to the mean SUV of the aorta for both patient and phantom images.

## 4. DISCUSSION

This work introduces PixelPrint^PET^, a 3D printing approach for generating PET phantoms with realistic organ geometry and activity distribution without the need for complex filling procedures or extensive post-processing. Compared with existing 3D printing-based methods, PixelPrint^PET^ offers several advantages, as described below:

1. PixelPrint^PET^ enables direct printing from patient images or user-defined models without requiring segmentation, thereby simplifying data processing. Previous anatomically based 3D-printed phantoms often rely on atlas data or segmentation from other modalities^8,9,12^, which lack patient-specific radiotracer absorption detail and are constrained by data availability. Other approaches, such as segmenting lesions from PET images, are limited by challenges in edge delineation and may result in loss of structural information^10,11^. In contrast, PixelPrint^PET^ allows straightforward replication of both patient-specific anatomies and standardized phantoms, such as NEMA designs.
2. PixelPrint^PET^ produces voxel-wise variations in activity, enabling a realistic representation of signal intensity and texture. Contrast variation was achieved through differences in porous structure density, similar to the approach used in petInsert^26^, which generates single fixed contrast inserts for 4D PET/CT. In contrast, PixelPrint^PET^ provides greater flexibility in tissue variety coverage, contrast modulation, and structural representation. In comparison, many existing methods assume homogeneous activity within each region and achieve contrast by filling compartments with different activity concentrations, which limits spatial heterogeneity^9,10,12^.
3. The absence of physical boundaries between regions eliminates wall-induced partial volume effects and improves anatomical fidelity.
4. The filling procedure is simple and robust, requiring only a single activity concentration, with contrast differences generated by the internal printed structure. This reduces preparation complexity and minimizes operator exposure to radioactive materials.
5. The phantoms are reusable and safe to store, unlike approaches that incorporate radioactive ink or resin directly into the printed object^8,27–29^. After radioactive decay, the solution can be removed, and the phantom can be cleaned and reused for subsequent experiments. No specialized imaging protocols or post-processing steps are required.
6. The method demonstrates good reproducibility, as identical phantoms can be fabricated and yield consistent results across repeated scans at different count levels, with variations below 5%.

Along with these advantages, several limitations should be noted. Because the phantom is immersed in the radioactive solution, the background signal outside of the phantom is elevated compared to in vivo conditions, but does not affect quantification of internal structures. Future work will focus on improving the filling setup, for example, by using flexible or custom-shaped containers that better conform to the phantom geometry and reduce background activity.

Partial volume effects remain a significant factor influencing quantitative accuracy. As observed in the CRC analysis of the contrast recovery phantom and in the brain phantom results, smaller or edge-adjacent regions show reduced contrast recovery, and the complex geometry of brain structures further amplifies these effects. A similar effect was observed in the thorax phantom, where the myocardium, bordered by the low-signal lung region, showed lower signal than expected. Since PVE originates from the inherent resolution of the PET scanner rather than the phantom, simulated images of the digital phantom generated with GATE were used as a reference. The good agreement between simulated and measured images confirms that the observed deviations reflect scanner resolution, which can be quantified and corrected. In addition, the use of patient-derived input data may introduce physiological noise, such as motion, and can lead to blurring at region boundaries in the printed phantom. Using high-quality input images helps minimize these effects.

The choice of background region also affects quantitative metrics such as TBR. Variability in the background signal can introduce bias across measurements, highlighting the need for more robust and standardized evaluation strategies. In addition, regions with high infill ratios, corresponding to lower PET signal, showed a systematic high positive bias in measured signal compared to expected values, as shown in CSF in the brain phantom and lung in the thorax phantom. This may be related to printing instability at larger line widths or the presence of small, trapped air bubbles not visualized on CT.

Future developments will focus on extending the method to incorporate filaments with different attenuation properties and multi-material printing techniques. For example, filaments containing calcium could be used to represent high-attenuation structures such as bone or vascular calcifications^10,11,28^. Using the current methodology, structures that have high tracer uptake must have low inflow ratios, which in turn correlates to low density on CT, creating a mismatch between uptake and density that does not necessarily reflect reality. The use of new materials could enable simultaneous and more accurate representation of both PET activity and CT density and could facilitate integration with spectral CT to provide additional information. Such improvements would further enhance the accuracy and utility of PixelPrint^PET^ phantoms for multimodal imaging applications.

## 5. CONCLUSION

In this study, we introduced PixelPrint^PET^, a 3D printing–based method for manufacturing PET/CT phantoms with anatomically realistic geometry and spatially heterogeneous radiotracer distributions. By enabling voxel-level spatial control of fillable volume, the method generates phantoms without physical boundaries between regions while preserving realistic image texture and contrast. The approach supports direct conversion from patient images without segmentation and provides a simple and robust workflow for fabrication, filling, and imaging, while demonstrating accurate signal representation, flexibility across tissue types and contrast levels, and strong reproducibility. Together, these capabilities establish PixelPrint^PET^ as a practical and scalable solution for creating realistic imaging phantoms, with the potential to accelerate the development, validation, and clinical translation of PET/CT technologies.

## Data Availability

All data produced in the present study are available upon reasonable request to the authors.

